# Identification of Outcome-Oriented Progression Subtypes from Mild Cognitive Impairment to Alzheimer’s Disease Using Electronic Health Records

**DOI:** 10.1101/2023.07.27.23293270

**Authors:** Jie Xu, Rui Yin, Yu Huang, Hannah Gao, Yonghui Wu, Jingchuan Guo, Glenn E Smith, Steven T DeKosky, Fei Wang, Yi Guo, Jiang Bian

**Affiliations:** Department of Health Outcomes &Biomedical Informatics, University of Florida, Gainesville, FL, USA; Hamilton Southeastern High School, Fishers, Indiana, IN, USA; Department of Pharmaceutical Outcomes & Policy, University of Florida, Gainesville, FL, USA; Department of Clinical and Health Psychology, University of Florida, Gainesville, FL, USA; Department of Neurology, College of Medicine, University of Florida, Gainesville, FL, USA; Department of Population Health Sciences, Weill Cornell Medicine, New York, NY, USA

## Abstract

Alzheimer’s disease (AD) is a complex heterogeneous neurodegenerative disease that requires an in-depth understanding of its progression pathways and contributing factors to develop effective risk stratification and prevention strategies. In this study, we proposed an outcome-oriented model to identify progression pathways from mild cognitive impairment (MCI) to AD using electronic health records (EHRs) from the OneFlorida+ Clinical Research Consortium. To achieve this, we employed the long short-term memory (LSTM) network to extract relevant information from the sequential records of each patient. The hierarchical agglomerative clustering was then applied to the learned representation to group patients based on their progression subtypes. Our approach identified multiple progression pathways, each of which represented distinct patterns of disease progression from MCI to AD. These pathways can serve as a valuable resource for researchers to understand the factors influencing AD progression and to develop personalized interventions to delay or prevent the onset of the disease.

## Introduction

Alzheimer’s Disease (AD), the most common type of dementia, is a progressive, irreversible, and heterogeneous neurodegenerative disorder, affecting millions of people worldwide.^1^ 6.7 million Americans are living with AD dementia,^2^ and the number is projected to reach 13.8 million by 2050.^3^ Such many AD-related populations will place a tremendous burden on patients, their families, the healthcare system, and even society. Mild cognitive impairment (MCI) is a translational intermediate state between normal cognitive function and dementia.^4,5^ It is a heterogeneous condition characterized by diverse cognitive profiles and clinical progression patterns, making it difficult to predict the outcomes and progression patterns for patients with MCI.^6^ The management of AD has identified MCI as a crucial target for both prognosis and therapy.^2^ However, not all MCI patients will convert to AD, and approximately 10% to 20% of individuals with MCI will advance to AD within one year, while the remaining individuals who do not progress to AD may either experience other types of dementia or maintain stability.^7,8^ It remains unclear whether MCI-to-AD patients experience a consistent rate of decline throughout the progression of their disease or if their trajectories change over time, possibly due to endogenous or exogenous factors.^9,10^ Therefore, understanding the progression of those individuals progressing from MCI to AD and identifying early diagnostic markers are of increasing clinical importance, which is essential to develop effective therapies and improve the quality of patients’ life.

Many recent studies have combined heterogeneous data sources to study the progression of MCI-to-AD through a combination of clinical markers and biomarkers such as magnetic resonance imaging (MRI)-based features,^11^ positron emission tomography,^12^ and cerebrospinal fluid.^13^ These clinical markers and biomarkers can be used to monitor the disease and to track changes in the brain over time,^14^ which have allowed researchers to better understand the underlying pathology of AD and to identify potential targets for treatments. However, previous studies usually utilized a restricted range of features from only a few modalities, with a particular emphasis on neuroimaging data, and some of the data sources like biomarkers typically require more invasive procedures or specialized equipment to obtain.

Numerous studies have attempted to classify and predict AD progression through diverse data types, while there remains a substantial disparity between research outcomes and the practical application of these systems in routine clinical practice.^15^ The increasingly available Electronic Health Records (EHRs) have made it possible to identify the patterns and subtypes of AD progression using data-driven approaches reflecting real-world clinical practice,^16^ as they contain a wide variety of critical health events of patients collected through routine care, including diagnostic codes, medication use, laboratory test results, and other relevant clinical data. These new data sources may provide new insights into the study of the underlying heterogeneity of AD and dementia. For example, a previous study used hierarchical clustering on longitudinal EHRs to computationally generate probable AD and related dementia sub-phenotypes with machine learning approaches.^17^ Four potential sub-phenotypes of AD and dementia were identified, which showed a correlation with mental health conditions and cardiovascular diseases. These subtypes exhibited significant differences in patient demographics, comorbidities, and treatments.^17^ Another study examined the natural progression of cognitive decline in patients with AD using EHR data.^18^ The results suggested that the rate of cognitive decline varied widely among patients, with some patients experiencing rapid decline and others showing slower rates of decline over time. Moreover, an unsupervised framework was developed with a representation learning model to analyze EHRs from the Mount Sinai Health System to identify subtypes characterized by varying degrees of dementia symptoms.^19^ This framework enables the creation of patient representations that facilitate large-scale patient stratification in a precise and efficient manner. More recently, a multi-modal AD progress prediction model was presented that incorporates both EHR and MRI data to classify into three different stages. It trained a deep auto-encoder to extract features from EHR data, and ResNet and 3D U-Net for MRI image data, followed by an entropy-based weighted sum classification method to combine the results from each modality and generate a final prediction.^20^

The traditional approach to clustering involves grouping patients based on their static or longitudinal covariates in an unsupervised manner.^21^ However, this approach does not take the observed outcomes of the patients into consideration, such as the onset of comorbidities, and adverse events. Ideally, the identified clusters (or subtypes) would contain patients not only of similar characteristics but also similar disease outcomes; and without the “*predicted*” outcome information, disease subtypes obtained from this type of clustering is of limited prognostic use to clinicians and patients. The growing interest in the use of outcome-oriented disease subtyping has captured much attention to classify individuals into subgroups based on their response to a particular progression or treatment.^22^ This approach is often used to help develop personalize treatment options. For example, Eshaghi et al. identified multiple sclerosis subtypes using unsupervised machine learning and MRI data.^23^ According to the findings, the subtypes identified through MRI can be used to predict the progression of disability and treatment response in patients with multiple sclerosis. These subtypes can also be utilized to categorize patients into specific groups for interventional trials. In sum, the outcome-oriented disease subtyping shows promise in identifying patient subgroups that share similar disease outcomes and responses to treatment and can be applied to a wide range of diseases.

The objective of this paper is to develop a computational approach that can identify outcome-oriented progression pathways from MCI to AD using large collections of EHRs. To achieve this goal, we employed machine learning techniques that can capture the heterogeneity of MCI to AD progression subtypes over time. Specifically, we utilized a deep learning approach based on the Long Short-Term Memory (LSTM) architecture^24^ to predict the onset of AD using data pf MCI patients, and learned representations of subsequences extracted from patients’ EHRs. Each subsequence is a temporal trajectory which captures the sequences of clinical measurements or events in EHRs. Hierarchical clustering techniques^25^ were then applied to group the representations into different clusters (i.e., patients’ states in this study). After linking the different states for each patient, we observed several merged progression subtypes (i.e., progression patterns or pathways). The proposed approach was evaluated on two large EHR datasets (referred to as Site A and Site B within the context of the study) randomly selected from the OneFlorida+ Clinical Research Consortium, and the results demonstrated the existence of specific progression pathways leading to AD. By leveraging EHR data and machine learning, our approach can facilitate earlier diagnosis and intervention for AD, ultimately improving the quality of life for patients and their families. **Figure 1** illustrate the framework for identifying MCI to AD progression subtypes.

**Figure 1.**
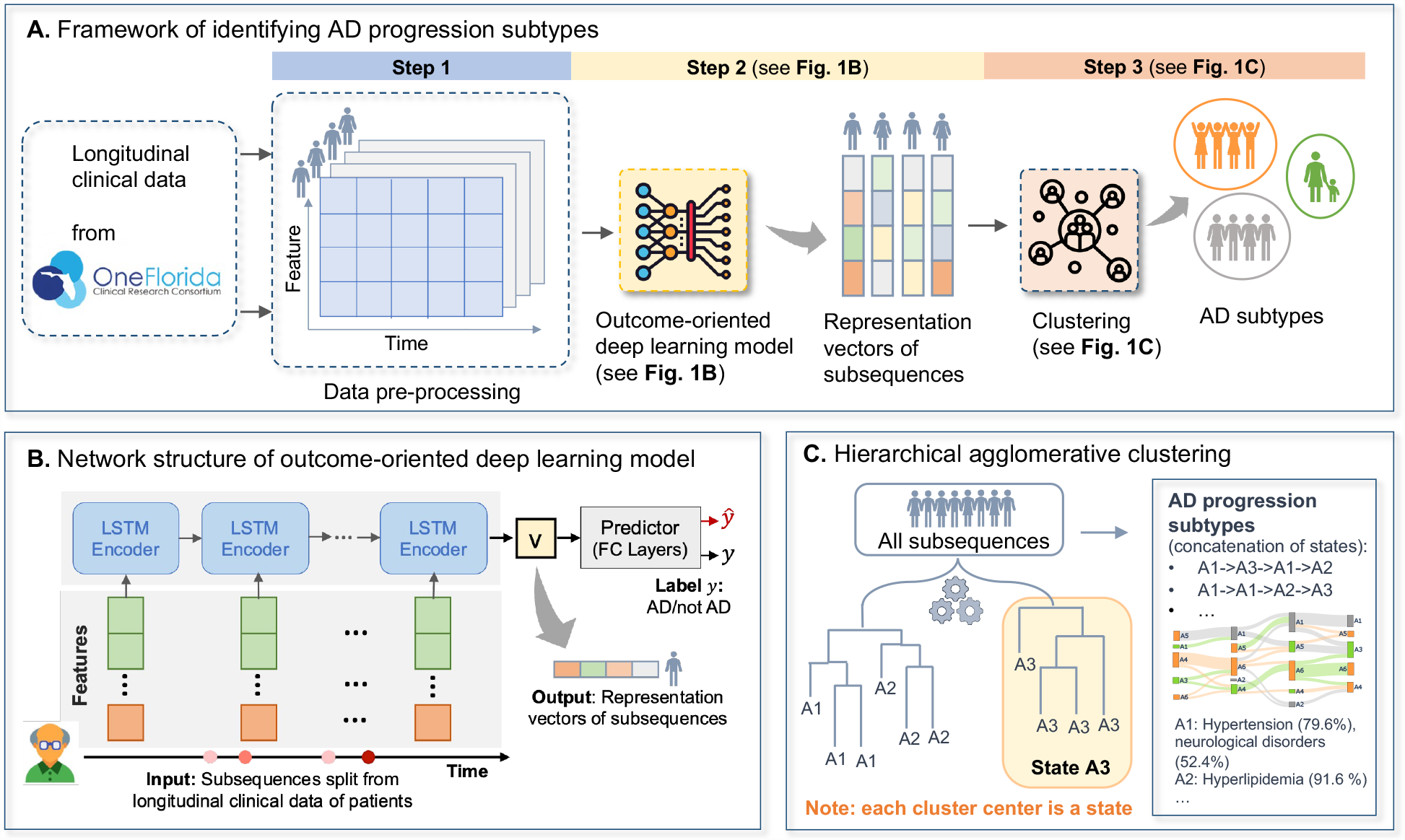
Illustration of the framework: (a) Framework of identifying MCI to AD progression subtypes; (b) Network structure of outcome-oriented deep learning model; and (c) Illustration of hierarchical agglomerative clustering. A patient’ longitudinal EHRs (i.e., a sequence) were divided into several subsequences. Subsequences were fed as input to the deep learning model to learn their representations. The learned representations were then subjected to hierarchical agglomerative clustering to derive clusters (i.e., states). By concatenating the states for each patient, we observed the progression subtypes.

## Methods

### Data source and study population

The study used large collections of EHR data from the OneFlorida+ Clinical Research Consortium, a clinical research network contributing to the national Patient-Centered Clinical Research Network (PCORnet). The OneFlorida+ network is a collaboration among 14 health organizations, including academic health centers and community health systems and clinics, covering 20 million patients from Florida (∼16.8 million), Georgia (∼2.1 million), and Alabama (∼1 million). The OneFlorida+ data, which followed the PCORnet Common Data Model (CDM), contained detailed patient information such as patient demographics, enrollment status, vital signs, conditions, encounters, diagnoses, procedures, prescribing, dispensing, and lab results. To create the study cohorts, we randomly selected two sites, referred to as site A and site B in our study, to consider between-site patient population heterogeneity. The study has been approved by the University of Florida Institutional Review Board (protocol no. IRB202202820).

Patients were eligible for the study if: (1) they had a diagnosis of MCI after January 2012; and (2) they were 50 years or older at the time of MCI diagnosis. The diagnosis of MCI was identified using ICD codes, specifically ICD 9 codes 331.83 and 294.9, and ICD 10 codes G31.84 and F09. To identify patients with AD among the MCI cohort, we used ICD codes, including ICD 9 code 331.0 and ICD 10 codes G30.*. Patients who had received a diagnosis of AD before their MCI diagnosis were excluded from the study.

### Construct temporal trajectory using longitudinal EHRs

To construct the temporal trajectories which are sequences of clinical measurements or events in EHRs, such as lab results, vital signs, diagnoses, or treatments, we aggregated the patient’s EHRs into specific time intervals, as there may be several clinical events that occur during the patient’s timeline. **Figure 2** illustrates how the AD trajectory is constructed using a patient’s EHRs, considering data before AD for the patients that converted to AD and all data until the end of the timeline for non-AD patients. The time window was calculated from the MCI onset date based on the first occurrence of MCI-related diagnostic codes (i.e., as the index date). As shown in **Figure 2**, relevant EHR data for each patient will be aggregated in 3-month blocks (i.e., window sizes) into a set of vectors.

**Figure 2.**
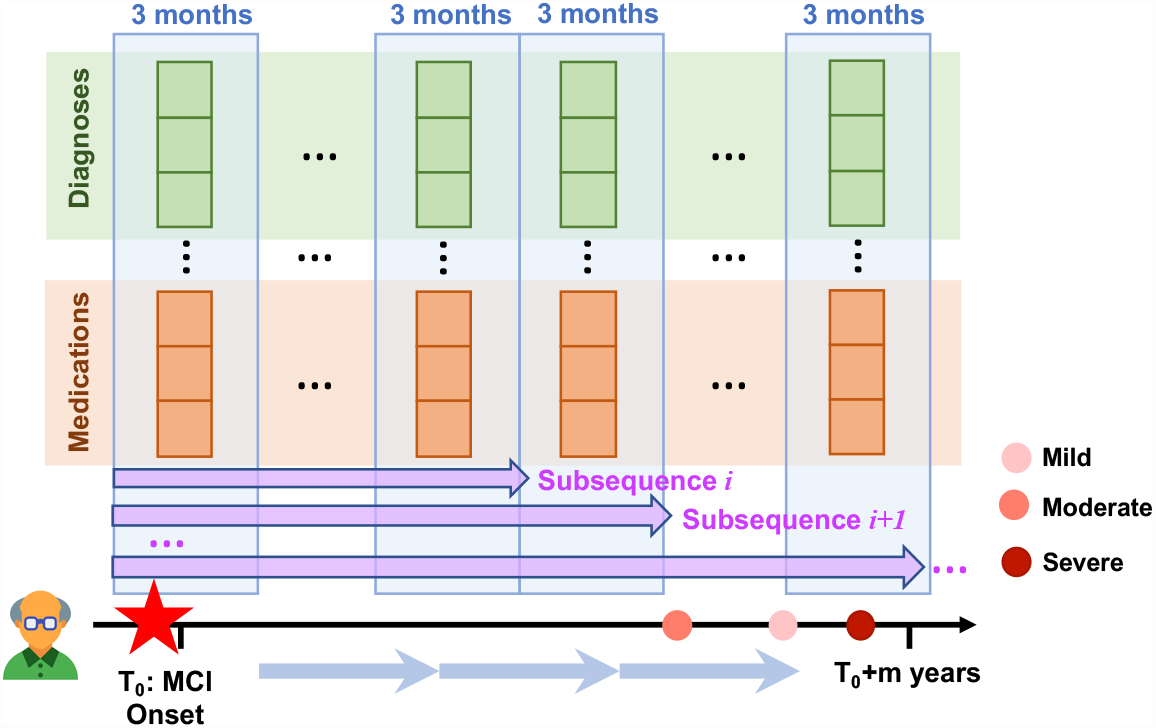
The AD temporal trajectory in EHRs.

To ensure that the data for each patient had a sufficient duration for learning the temporal representation, we imposed two additional inclusion criteria. First, patients were required to have at least one year of data before and after the index date. Second, patients were required to have a conversion time to AD of more than half a year. Each vector corresponds to a specific event type, such as diagnosis, medication, etc., based on discrete structured EHR data. Age was discretized using uniform-sized bins, and one-hot encoding was used to encode age, gender, and race variables.^27^ The diagnosis codes were mapped to Phecode, which is designed to support phenome-wide association studies (PheWAS) in EHRs. Drug codes, such as National Drug Codes (NDC) and RxNorm, were mapped to the third level of the Anatomical Therapeutic Chemical (ATC) Classification System. Finally, all features, including diagnosis and medication, were concatenated to represent each patient as a binary vector.

In addition, to model the progression pattern, we split each patient into multiple subsequences. All subsequences started from the index date (i.e., MCI onset date), and every 3 months served as a time point. A new subsequence was created every 6 months until the data reached its maximum length of a patient’s EHRs. Each subsequence was treated as an independent data sample and fed into the model. Mathematically, the trajectory of the *n*-th patient’ *l*-th subsequence can be represented as 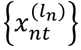, where *t* ∈ {1,2, …, *T*_*n*_} is the timestamp index such that *t* = 1 means the first 3 months after the index date, *l* ∈ {1,2, …, *l*_*n*_} is the index of subsequences split from *n*-th patient’s EHRs, and *x* is the binary vector (e.g., diagnoses, medications, etc.) constructed using the data before time point *t*. Finally, we have 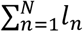 subsequences with *N* equals to the number of patients in the study cohorts.

### Deriving outcome-oriented temporal representation using outcome-oriented LSTM

After constructing the temporal trajectory of each sample, outcome-oriented LSTM is then applied to learn representations of the subsequences of a patient.^28^ **Figure 1B** shows the outcome-oriented LSTM model’s network structure, which comprises two components: an LSTM encoder and a predictor. The LSTM encoder cell takes in the multivariate time-series subsequence of each patient as input and generates a hidden state, which represents the patient’s state over time. By using “*memory cells*,” the LSTM can store historical information for extended periods, making it an excellent candidate for modeling disease progression based on longitudinal clinical data from patients.^29^ The learned vector representation of a patient’s subsequence is then passed on to the predictor, which attempts to predict whether the patient has AD or not. The model was trained by minimizing the difference between the actual and predicted labels. After the training process, a learned vector representation is obtained for each subsequence, considering the AD outcomes.

### Deriving progression subtypes with hierarchical agglomerative clustering

Once the temporal representation of each subsequence has been learned, the next step is to utilize clustering techniques to identify clusters (or states) of subsequences that exhibit similar characteristics. **Figure 1C** illustrates the clustering process using hierarchical clustering^25^. Each subsequence is initially treated as a separate cluster, and then the two closest clusters are merged repeatedly until all clusters are merged. The grouping of subsequences is based on the similarity of temporal representation vectors, which was learned in the previous step. The final goal is to obtain clusters of subsequences with distinct features, where each cluster center represents a state. Once the clusters of subsequences are identified, the states for each patient at different time points can be determined by the cluster centers of the corresponding subsequences. The trajectory pattern of a patient can be represented by concatenating different states which are the corresponding clustering centers of the subsequences extracted from the patient’ EHR, indicating the progression from one state to another state. For example, if a patient’s medical records were split into four subsequences, and the clustering algorithm identified three states (e.g., A1, A2, A3), then the trajectory pattern of that patient might be “*A1->A3->A1->A2*”, as illustrated in **Figure 1C**. After merging the patients with similar trajectory patterns, we could observe the final progression subtypes.

## Results

**Table 1** presents the characteristics of the study cohorts, which consist of patients from both Site A and Site B. As shown in the table, a higher percentage of patients from Site B transitioned to AD (i.e., 12.6%), and the average duration of their transition was longer than that of Site A (i.e., 907.2 days). Moreover, patients at Site B had an average age of 76.6 years, indicating that this group was relatively older than patients at Site A, whose average age was 70. Additionally, Site A had a greater proportion of Hispanic patients, accounting for 38.8% of the total patient population, while Site B had fewer Hispanic patients and a higher proportion of White patients than Site A.

**Table 1.**
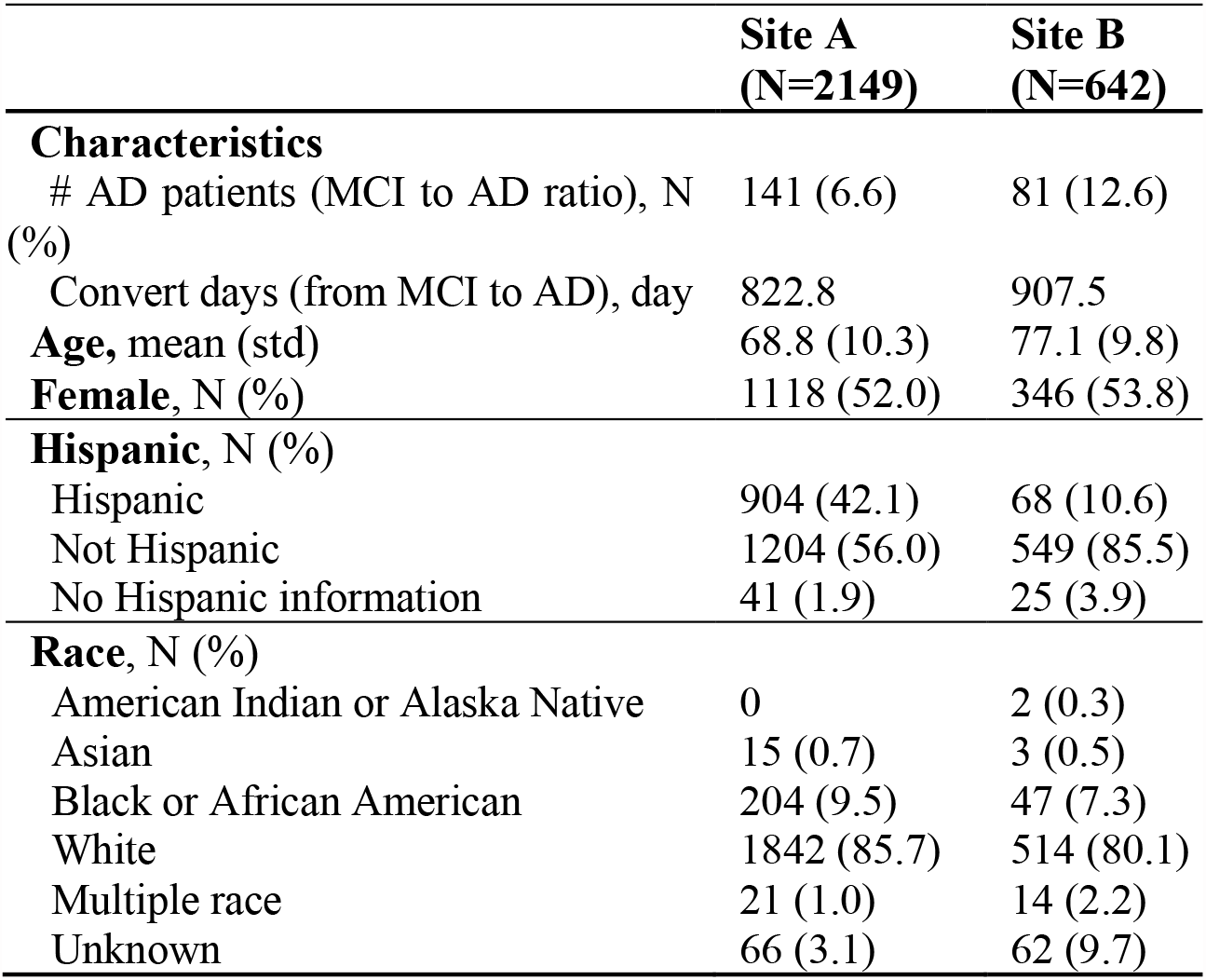
Characteristics of the study cohort.

From site A, six clusters (i.e., states) were derived by analyzing dendrogram: state A0 (N=1737; 80.83%); state A1 (N=183; 8.52%); state A2 (N=89; 4.14%); state A3 (N=79; 3.68%); state A4 (N=46; 2.14%); and state A5 (N=15; 6.98%). The clustering results were visualized in **Figure 3(a)**, where the left figure depicted the uniform manifold approximation and projection (UMAP)^30^ of the six states. To gain insights into the features that differentiated these states, the patient percentages of the top 20 features for each state were displayed in the left figure of Figure 3(c). The analysis revealed that essential hypertension was the most prevalent disease among all the clusters. Moreover, state A0 included patients with fewer comorbidities, while state A3 comprised patients with more comorbidities. To further compare the differences between these states, statistical analysis was performed using Chi-square tests to examine the significant differences between two states. The p-values of these statistical tests are illustrated in the right part of Figure 3(c). It was observed that state A0 and A1 were significantly different from one another, except for primary open angle glaucoma. This difference exhibited between A0 and A4 could be crucial in understanding the progression from MCI to AD as this pathway is directly towards to AD.

**Figure 3.**
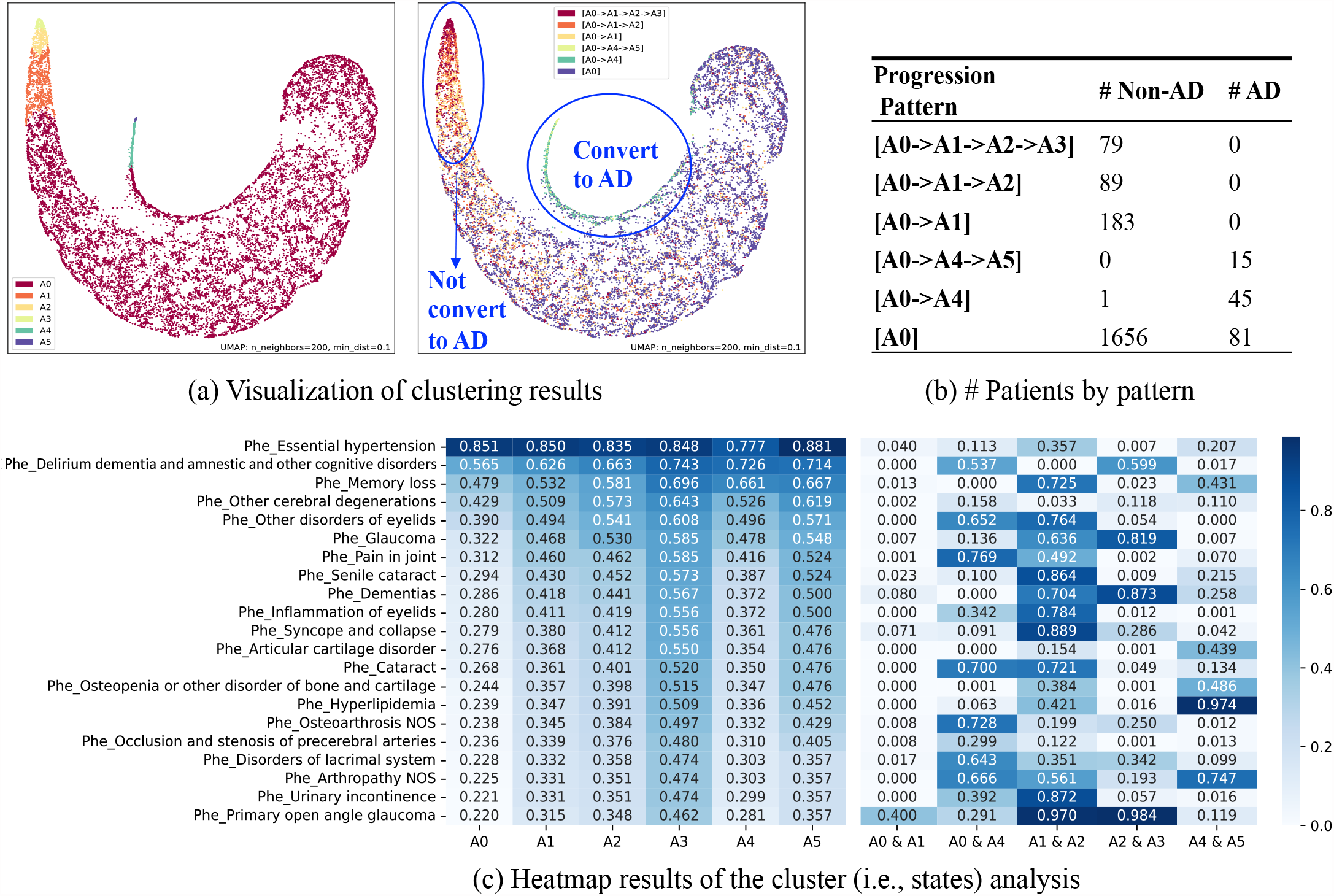
Clustering results of site A. (a) Visualization of clustering results; (b) Number of patients by progression pattern (i.e., subtype); and (c) Heatmap results of the cluster (i.e., states) analysis.

The analysis of the states of the subsequences split from each patient resulted in the identification of six distinct progression subtypes: (1) A0->A1->A2->A3, (2) A0->A1->A2, (3) A0->A1, (4) A0->A4->A5, (5) A0->A4, and (6) A0. **Figure 3(b)** shows that patients who converted from state A0 to state A1 did not progress to AD. However, most patients who converted to state A4 eventually progressed to AD. And A0 and A4 are significantly different in memory loss, dementias, articular cartilage disorder. Further analysis revealed that the average time taken for patients to progress to AD in progression subtype (4) A0->A4->A5 was 1391.1 days, whereas the average time taken to progress to AD in subtype (5) A0->A4 alone was 1117.2 days. For patients who exhibited a progression pattern to AD in pattern (6) A0, the average time taken was relatively short, with an average of 554.1 days. Nevertheless, in some cases, there may not be enough longitudinal data to observe a clear progression pattern from state A0 for patents in pattern (6) A0.

For subsequences split from patients of site B, five clusters (i.e., states) were derived based on dendrogram: state B0 (N=1737; 80.83%); state B1 (N=183; 8.52%); state B2 (N=89; 4.14%); state B3 (N=79; 3.68%); state B4 (N=46; 2.14%); and state A5 (N=15; 6.98%). The clustering states are shown in **Figure 4**. Similar to site A, to check the significant difference between the states, a chi-square test was applied, and the p-value results are presented in the right figure of Figure 4(c). The patient percentage of top features among each state is also depicted in the left figure of Figure 4(c).

**Figure 4.**
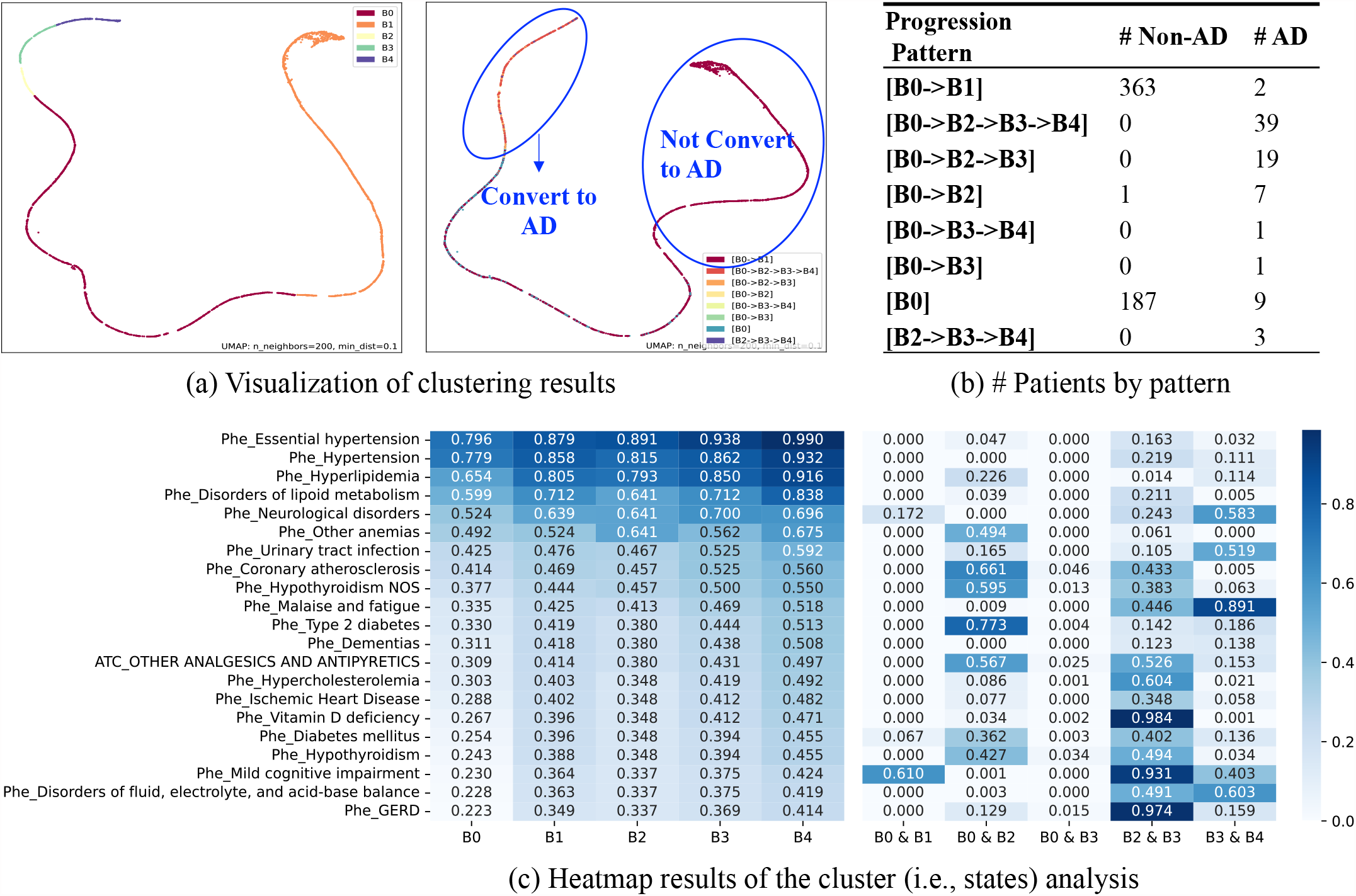
Clustering results of site B. (a) Visualization of clustering results; (b) Number of patients by progression pattern (i.e., subtype); and (c) Heatmap results of the cluster (i.e., states) analysis.

By linking the states of the subsequences from patients at site B, eight progression subtypes were identified: (1) B0->B1, with an average conversion time of 675.5 days to AD, (2) B0->B2->B3->B4, with an average conversion time of 1179.6 days to AD, (3) B0->B2->B3, with conversion time of 721.3 days to AD, (4) B0->B2, with conversion time of 574.9 days, (5) B0->B3->B4, with conversion time of 1704 days, (6) B0->B3, with conversion time of 245 days, (7) B0, with conversion time of 307.4 days, and (8) B2->B3->B4, with conversion time of 1238.3 days to AD. Figure 4(b) shows the conversion pattern of patients from state B0 to other states. The results suggest that if patients convert from state B0 to state B1, most of them didn’t convert to AD. For patients who convert to state B2, most of them progress to AD. The reason for not being able to observe the next state for patients in subtype (7) B0 could be attributed to inadequate data for some patients, which prevented the observation of the subsequent state.

## Discussion and Conclusion

We developed a machine learning approach using longitudinal EHRs to identify distinct progression pathways leading to AD from MCI. LSTM model and hierarchical clustering techniques were used to group trajectory patterns. The approach was evaluated on two datasets from two health system sites randomly selected from the OneFlorida+ network. In both datasets, we were able to identify multiple subtypes of patients with distinct progression patterns from MCI to AD. These patterns suggest that MCI is not a uniform disease state and that different subtypes of MCI patients may exist, each with unique progression trajectories towards AD. By better understanding the distinct subtypes of patients and their disease progression patterns, we may be able to develop more personalized and effective treatments for individuals with MCI, slowing down or preventing from their progression to AD. This approach highlights the potential of leveraging EHR data and machine learning to facilitate earlier diagnosis and intervention for AD, ultimately improving the quality of life for patients and their families.

The study cohorts consisted of patients from two heterogenous sites from the OneFlorida+ network, Site A and Site B, with different characteristics. The results showed that a higher proportion of patients from Site B transitioned to AD but in average has a longer conversion time than Site A patients. Patients at Site B were also found to be older on average than Site A patients. This finding further confirms the significance of age in the development and progression of AD, as older individuals were found to be more susceptible to the disease.^31^ Our results demonstrated the significant differences in the population and their data distribution across different sites. Therefore, it is crucial to consider the potential impact of such differences (i.e., between-site heterogeneity) when analyzing EHRs from different populations (e.g., geographic regions and population composition).

Our analysis of both data sources revealed that there are two main distinct progression pathways that can be identified. The first pathway involves patients who initially convert to a specific state (i.e., A4 in Site A and B2 in site B), which ultimately leads to a conversion to AD. Specifically, both A0 and A4, B0 and B2 are significantly different in features including memory loss, dementias, nonspecific abnormal findings on radiological and other examination of skull and head, and other and unspecified coagulation defects. In contrast, the second pathway involves patients who convert to a different state (i.e., A1 in Site A and B1 in Site B), and these patients do not convert to AD. These findings suggest that the progression of the disease may vary depending on the initial state that patients convert to. However, more research is required to validate these findings and investigate the mechanisms driving the different progression pathways. Such an analysis could provide valuable insights into the underlying patterns of diseases and comorbidities that are associated with the progression of a patient from one state to another. This could help in identifying the critical factors that influence disease progression and developing effective strategies for managing and treating patients in each state.

The study is subject to several limitations. Firstly, the data used was only from a specific region, and this could limit the generalizability of the study’s findings to other populations with distinct demographics, healthcare systems, and policies. Secondly, the study only considered the presence or absence of AD as the outcome label for studying progression patterns. This approach may not provide sufficient detail about the severity of the disease, as it overlooks critical indicators of disease progression. To gain a more nuanced understanding of AD progression patterns, various factors reflecting disease severity must be considered. These factors encompass cognitive decline, behavioral changes, physical deterioration, and medical complications, which can be utilized to evaluate the severity of AD and facilitate a comprehensive analysis of disease progression patterns. Thirdly, the study relied on EHRs, which have advantages over paper-based systems, but may not capture all relevant patient data and may not be standardized across different healthcare providers.^32^ Moreover, the use of EHRs may overlook critical factors that impact the diagnostic process, such as clinician preferences, access to diagnostic tools, and administrative rules, which could introduce bias into the study’s results. Additionally, EHRs may contain errors or glitches, potentially impacting the accuracy of the collected data. These limitations highlight the importance of caution when interpreting the study’s findings and emphasize the need for future research to consider these limitations to improve the accuracy and generalizability of findings.

The future work of this study will concentrate on developing validated phenotyping algorithms and predictive models that can identify the critical features responsible for patient transitions between different states using the states as the outcome label. In addition to this, the study will also apply causal inference techniques to determine the impact of each risk factor on AD progression.^33^ Furthermore, to enrich and normalize the progression subtypes across various healthcare institutions, we will also apply federated learning, a privacy-preserving machine learning technique that avoids the aggregation of raw clinical data locally across different institutions.^34^ This will allow the researchers to combine the data from different institutions while ensuring patient privacy is maintained. By combining these predictive models with causal inference analyses, we hope to gain a more complete understanding of the underlying mechanisms that drive Alzheimer’s disease and identify strategies for early diagnosis and prevention.

## Data Availability

The dataset used during the current study is a HIPAA limited data set, which requires a data use agreement with the OneFlorida+ clinical research consortium, https://onefloridaconsortium.org/. Request of the data can be sent to the OneFlorida+.

## Acknowledgments

This work was partially supported by a grant from the Ed and Ethel Moore Alzheimer’s Disease Research Program of the Florida Department of Health (FL DOH #23A09) and grants (R01AG080624, R01AG080991, R01AG076234, and UL1TR001427) from the National Institutes of Health (NIH).

